# A Cluster-Randomized Trial of Hydroxychloroquine as Prevention of Covid-19 Transmission and Disease

**DOI:** 10.1101/2020.07.20.20157651

**Authors:** Oriol Mitjà, Maria Ubals, Marc Corbacho-Monné, Andrea Alemany, Clara Suñer, Cristian Tebe, Aurelio Tobias, Judith Peñafiel, Ester Ballana, Carla A. Pérez, Pol Admella, Nuria Riera-Martí, Pep Laporte, Jordi Mitja, Mireia Clua, Laia Bertran, Maria Sarquella, Sergi Gavilán, Jordi Ara, Josep M Argimon, Gabriel Cuatrecasas, Paz Cañadas, Aleix Elizalde-Torrent, Robert Fabregat, Magí Farré, Anna Forcada, Gemma Flores-Mateo, Cristina López, Esteve Muntada, Núria Nadal, Silvia Narejos, Aroa N Gil-Ortega, Nuria Prat, Jordi Puig, Carles Quiñones, Ferran Ramírez-Viaplana, Juliana Reyes- Urueña, Eva Riveira-Muñoz, Lidia Ruiz, Sergi Sanz, Alexis Sentis, Alba Sierra, César Velasco, Rosa Maria Vivanco-Hidalgo, Juani Zamora, on behalf of the BCN PEP-COV RESEARCH GROUP, Jordi Casabona, Martí Vall-Mayans, Camila G-Beiras, Bonaventura Clotet

## Abstract

**Background:** Current strategies for preventing severe acute respiratory syndrome coronavirus 2 (SARS-CoV-2) infections are limited to non-pharmacological interventions. Hydroxychloroquine (HCQ) has been proposed as a postexposure therapy to prevent Coronavirus disease 2019 (Covid-19) but definitive evidence is lacking.

**Methods:** We conducted an open-label, cluster-randomized trial including asymptomatic contacts exposed to a PCR-positive Covid-19 case in Catalonia, Spain. Clusters were randomized to receive no specific therapy (control arm) or HCQ 800mg once, followed by 400mg daily for 6 days (intervention arm). The primary outcome was PCR-confirmed symptomatic Covid-19 within 14 days. The secondary outcome was SARS-CoV-2 infection, either symptomatically compatible or a PCR-positive result regardless of symptoms. Adverse events (AEs) were assessed up to 28 days.

**Results:** The analysis included 2,314 healthy contacts of 672 Covid-19 index cases identified between Mar 17 and Apr 28, 2020. A total of 1,198 were randomly allocated to usual care and 1,116 to HCQ therapy. There was no significant difference in the primary outcome of PCR-confirmed, symptomatic Covid-19 disease (6.2% usual care vs. 5.7% HCQ; risk ratio 0.89 [95% confidence interval 0.54-1.46]), nor evidence of beneficial effects on prevention of SARS-CoV-2 transmission (17.8% usual care vs. 18.7% HCQ). The incidence of AEs was higher in the intervention arm than in the control arm (5.9% usual care vs 51.6% HCQ), but no treatment-related serious AEs were reported.

**Conclusions:** Postexposure therapy with HCQ did not prevent SARS-CoV-2 disease and infection in healthy individuals exposed to a PCR-positive case. Our findings do not support HCQ as postexposure prophylaxis for Covid-19.

**ClinicalTrials.gov registration number:** NCT04304053

## INTRODUCTION

Coronavirus 2019 disease (Covid-19) is a rapidly emerging infection caused by the severe acute respiratory syndrome coronavirus 2 (SARS-CoV-2). The rate of new cases among contacts (secondary attack rate) has been estimated as 10 to 15%.^1-4^ The current infection control strategy is based on social distancing and isolation of cases and contacts.^5^ The effectiveness of the latter depends on the promptness of the intervention, level of contact tracing, and level of isolation compliance.^6^ Unfortunately, real-world constraints for implementing full effective measures have resulted in SARS-CoV-2 spread in many countries.

Postexposure prophylaxis of healthy contacts is among the measures used for outbreak control of several infectious diseases, for example, in pandemic influenza.^7^ No agent is known to be effective in preventing SARS-CoV-2 infection or disease, but several drugs have shown antiviral activity in the laboratory, including the aminoquinolines hydroxychloroquine (HCQ) and chloroquine.^8^ In-vitro results showed that these drugs block the SARS-CoV-2 viral spread in cell cultures^9–11^ and that HCQ was more effective at impairing SARS-CoV-2 viral replication compared to chloroquine.^11^ To date, only one RCT has reported on HCQ for postexposure prophylaxis for Covid-19.^12^ However, concerns have been raised about the trial design, primarily because most participants were diagnosed with an influenza-like illness based on symptoms alone, and only 20% of their Covid-19 outcome was confirmed with PCR.

We investigated the efficacy and safety of HCQ to prevent secondary PCR-confirmed symptomatic Covid-19 (confirmed Covid-19) and SARS-CoV-2 infection in contacts exposed to a PCR-positive Covid-19 case during the outbreak in Catalonia, the region with the second highest number of Covid-19 cases in Spain.

## METHODS

### PARTICIPANTS

We included adult individuals ≥ 18 years of age with a recent history of close contact exposure to a PCR-confirmed Covid-19 case (i.e., > 15 minutes within two meters, up to seven days before enrolment) and absence of Covid-19-like symptoms on the two weeks preceding enrolment, as either a healthcare worker, a household contact, a nursing home worker or a nursing home resident. Contacts with Covid-19-like signs and symptoms at the time of the baseline visit were considered unpreventable Covid-19 events and were not enrolled in the study. All eligibility criteria are listed in the Supplementary Appendix.

### TRIAL DESIGN AND OVERSIGHT

This was an open-label, phase 3 cluster-randomized trial conducted from Mar 17 to Apr 28, 2020, during the Covid-19 outbreak, in three out of nine health administrative regions in Catalonia, Spain: *Catalunya central, Àmbit Metropolità Nord, and Barcelona Ciutat* (total target population 4,206,440 people; Fig. S1, Supplementary Appendix).

Study candidates were screened using the electronic registry of the Epidemiological Surveillance Emergency Service of Catalonia (SUVEC) of the Department of Health. During the Covid-19 outbreak in Catalonia, a public health ordinance required all patients who tested positive for Covid-19 in any of the designated diagnostic laboratories to be notified to the SUVEC.^13^

The study protocol and subsequent amendments, available at NEJM.org, were approved by the institutional review board of Hospital Germans Trias Pujol, and the Spanish Agency of Medicines and Medical Devices. All participants provided written informed consent.

### TRIAL PROCEDURES

Following a similar approach as the ring vaccination trial “Ebola Ça Suffit!”,^14^ we defined study clusters (called rings) of healthy individuals (contacts) epidemiologically linked to a PCR-positive Covid-19 case (index case). All contacts in a ring were simultaneously cluster-randomized (1:1) to either a control arm or an intervention arm. Randomization was performed remotely by a member of the study team not involved in participants’ enrollment. Following ring randomization, we verified the selection criteria of individual candidates and obtained informed consent for enrollment. The allocation was revealed to participants after providing written consent on day 1 (baseline). Participants allocated in the control arm received no treatment aside from usual care, whereas those in the intervention arm received HCQ (Dolquine^®^) 800 mg on day 1, followed by 400 mg once daily for six days. The dose and regimen of HCQ were chosen based on pharmacokinetic simulations to achieve plasma and lung concentrations above the SARS-CoV-2 half-maximal effective concentration observed in-vitro^11^ for 14 days (details provided in the Study Protocol).

By the time of trial conduct, quarantine was mandatory for all exposed contacts, according to the National Department of Health guidelines; hence the likelihood that a participant could be exposed to other cases was low. Covid-19 index cases that generated the rings were enrolled in a nested trial aimed at investigating the efficacy of early treatment with hydroxychloroquine as therapeutic intervention for Covid-19 outpatients. Laboratory technicians were unaware of participants’ treatment, treatment response, and previous PCR results during the entire follow-up period.

All contacts were visited at home or workplace on day 1 for medical exam, and baseline nasopharyngeal swab collection for SARS-CoV-2 RT-PCR test and viral load. Symptoms surveillance consisted of active monitoring by phone on days 3 and 7, and passive monitoring whenever the participant developed symptoms (i.e., participants were advised to call the research team). Participants who developed symptoms were visited the same day (unscheduled visit) by the outbreak field team for nasopharyngeal swab collection. All participants were visited at home on day 14 for nasopharyngeal swab collection, and finger-prick for IgM/IgG rapid test. Safety, medication adherence (i.e., treatment and number of doses taken), and crossover (i.e., unplanned conversion of control to intervention) were assessed using self-reports collected in telephone interviews on days 3, 7, and 28. Details on procedures performed at each visit and laboratory methods for SARS-CoV-2 identification and quantification (Fig. S2) are provided in the Supplementary Appendix.

### OUTCOMES

The primary outcome was the onset of a confirmed Covid-19 episode, defined as symptomatic illness (at least one of the following symptoms: fever, cough, difficulty breathing, myalgia, headache, sore throat, new olfactory and taste disorder(s), or diarrhea) and a positive SARS-CoV-2 RT-PCR test. The primary outcome was assessed in all asymptomatic individuals, irrespective of the PCR result; in a post hoc analysis, we explored the outcome in individuals with positive and negative PCR separately. Time-to-event was defined as the number of days from the date of randomization/exposure to the confirmed date of the onset of symptomatic illness.

The secondary outcome was the incidence of SARS-CoV-2 infection, defined as either the RT-PCR detection of SARS-CoV-2 in a nasopharyngeal specimen or the presence of any of the aforementioned symptoms compatible with Covid-19. The rationale for this outcome was to encompass definitions of Covid-19 used elsewhere^12,15^ and all possible viral dynamics. We, therefore, assumed that if clinical suspicion is high, infection should not be ruled out based on a negative PCR alone—particularly early in the course of infection.^15^ Participants who were hospitalized or died and whose hospital/vital records listed Covid-19 as the main diagnosis (including PCR confirmation) were also considered for the primary and secondary outcomes. We also measured serological positivity (IgM/IgG) of contacts at day 14. Safety outcomes included the frequency and severity of adverse events (AE), serious AE (SAE), and AE of special interest (e.g., cardiac) up to 28 days from treatment start. Causality was assessed by an external panel of pharmacovigilance consultants.

### STATISTICAL ANALYSIS

With an enrollment target of 95 clusters per trial group^16^ —15 participants per cluster and intraclass correlation of 1.0— the initial design yielded 90% power to detect a difference of 10% in the incidence, with expected incidence of 15% in the control arm. Owing to the limited information available by March 2020 regarding the cluster size and the incidence of Covid-19 after exposure, the protocol prespecified a sample-size re-estimation at the interim analysis. This re-estimation was aimed at maintaining the ability (80% power) to detect a reduction from 6.5% to 3% of the primary outcome, yielding 320 clusters per trial group with 3.5 participants per cluster.

The primary efficacy analysis was performed on the intention-to-treat (ITT) population, which included all randomized subjects with complete outcome data. We decided not to impute outcome data to participants with missing measurements because this approach would have biased the incidence of secondary Covid-19 events. Sensitivity analyses were performed with the per-protocol (PP) population in participants who completed the trial according to the protocol. The safety population included all participants who received any trial intervention, including usual care.

The cumulative incidence in primary, secondary, and safety outcomes was compared at the individual level using a binomial regression model with robust sandwich standard errors to account for clustering within rings.^17^ We defined a generalized linear model with a binomial distribution and a logarithm link function to estimate the relative risk (RR) as a measure of effect.^18^ The individual-level variables we adjusted for are age, gender, region, and time of exposure. We did additional pre-specified analyses to assess the consistency of treatment effects in subgroups defined according to the viral load of the contact at baseline, viral load of the index case, place of exposure, time of exposure to the index case. Survival curves by study groups on time-to-event outcomes were compared using a Cox proportional hazards model with a cluster-level frailty term to adjust for clustering.^19^ The significance threshold was set at a two-sided alpha value of 0.05, unless otherwise indicated, and all statistical analyses were conducted in R version 3.6.2.^20^

## RESULTS

### CHARACTERISTICS OF STUDY PARTICIPANTS

Between Mar 17 and Apr 28, 2020, we assessed 754 Covid-19 index cases for eligibility; 672 of them were selected for defining the corresponding clusters, which included 4,399 contacts (Fig. 1). 1,874 (42.6%) of the 4,399 contacts were not enrolled because of at least one exclusion criteria, including contacts presenting Covid-19-like symptoms before enrolment (n = 537). Additionally, 211 (8.4%) of 2,525 enrolled contacts were excluded from ITT analysis because of screening failure or missing PCR results on day 14, yielding an ITT population of 2,314 contacts. During follow-up, 64 participants had a protocol deviation regarding the intervention (PP population of 2,250 contacts).

**Figure 1.**
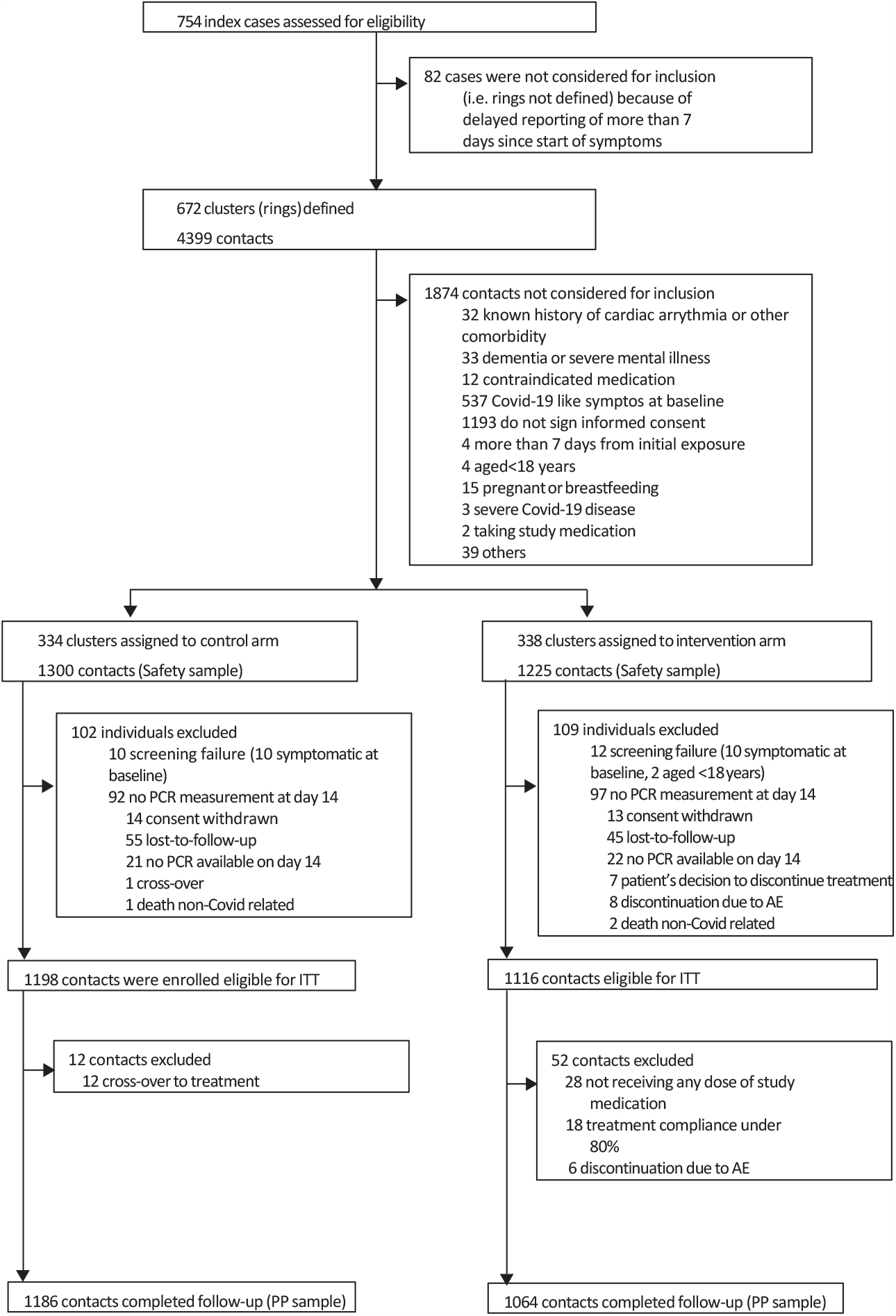
Flow diagram of individual selection and allocation. The safety population (n=2,497; 1,300 in the control arm and 1,197 in the intervention arm) included all individuals in the ITT population (except 28 not receiving any dose of study medication) plus 211 participants that received medication but were excluded from ITT because of screening-failure, or missing PCR results on Day 14.

The demographic, clinical, and epidemiological characteristics of participants at baseline were similar in the two study arms (Table 1, PP analysis in the Supplementary Appendix). The mean age of contacts was 48.6 years (SD 19.0) and the PCR test at baseline was negative in 87.8% of them (2,000 of 2,314). Overall, 55.6% of the participants (1,287 of 2,314) reported chronic health conditions. The median length from exposure to enrolment was 4.0 (IQR 3.0–6.0) days. The size of clusters was similar in both arms (median 2.0 vs. 2.0; *P* = 0.25). Exposure was predominantly from an index case with moderate-to-high viral load shedding (460 of 549 [83.8%] index cases with available viral load assessment). Health care workers and nursing home workers accounted for 60.3% (1,395) of the participants; 27.7% (640) were enrolled as household contacts, and 12.7% (293) as nursing home residents. Overall, 67.2% (1,555) of participants reported routine use of masks at the time of exposure, and 6.2% (144) of contacts continued to sleep in the same room as the index case.

**Table 1.**
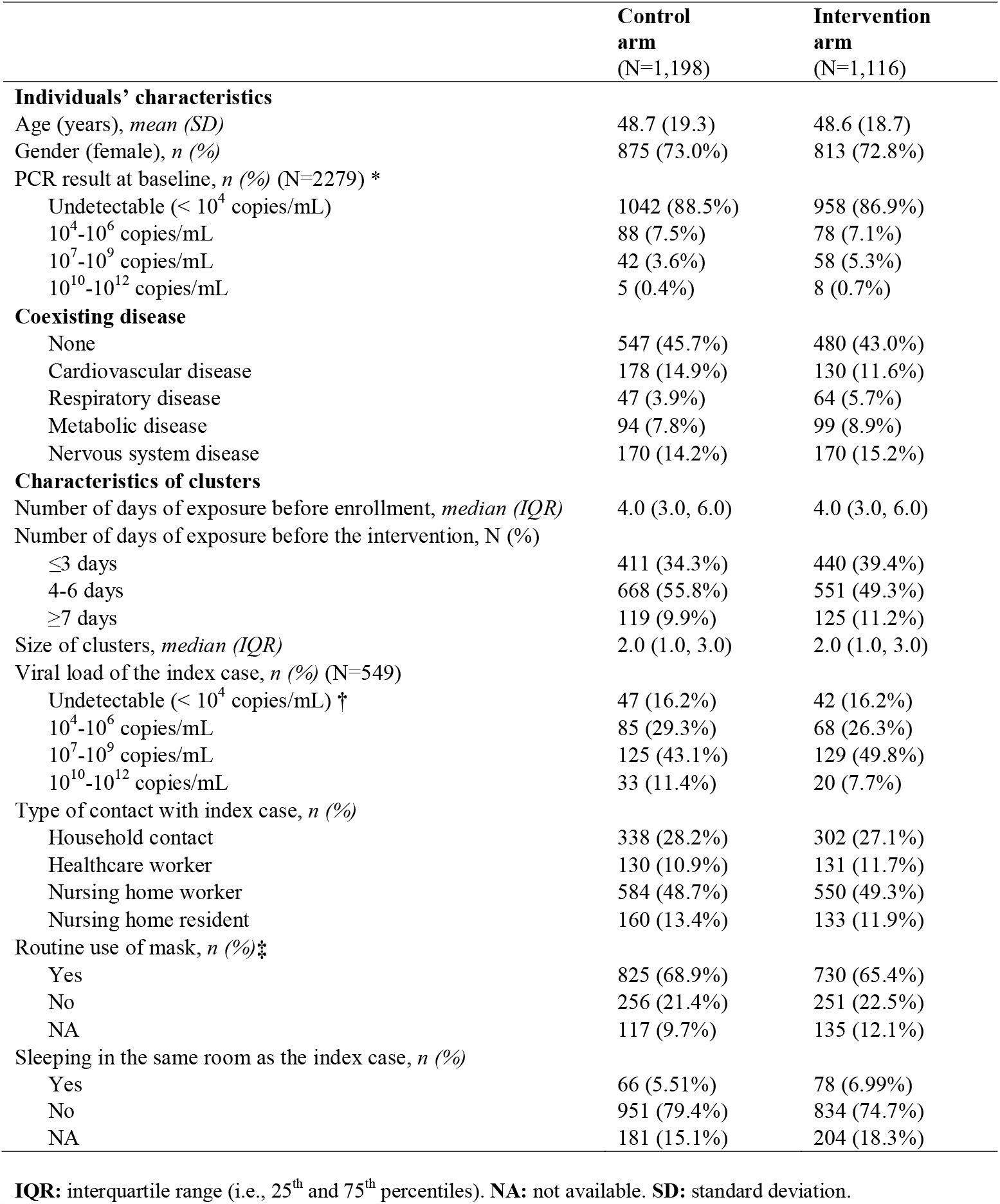
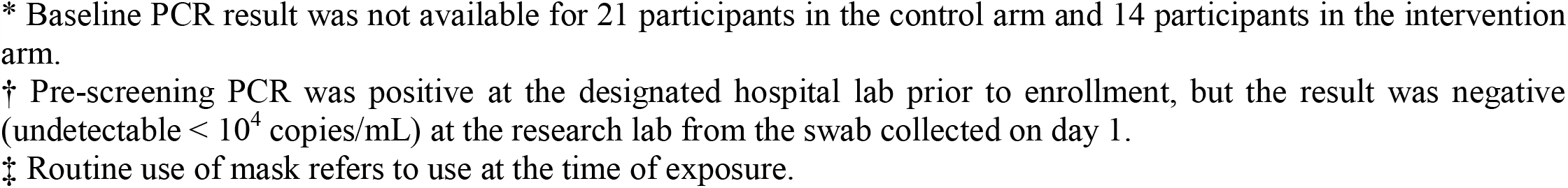
Baseline characteristics of study participants (contacts) included in the intention-to-treat population (N=2314).

|  | Control<br>arm<br>(N=1,198) | Intervention<br>arm<br>(N=1,116) |
| --- | --- | --- |
| **Individuals' characteristics** |  |  |
| Age (years), *mean (SD)* | 48.7 (19.3) | 48.6 (18.7) |
| Gender (female), *n (%)* | 875 (73.0%) | 813 (72.8%) |
| PCR result at baseline, *n (%)* (N=2279) * |  |  |
| Undetectable (< 10<sup>4</sup> copies/mL) | 1042 (88.5%) | 958 (86.9%) |
| 10<sup>4</sup>-10<sup>6</sup> copies/mL | 88 (7.5%) | 78 (7.1%) |
| 10<sup>7</sup>-10<sup>9</sup> copies/mL | 42 (3.6%) | 58 (5.3%) |
| 10<sup>10</sup>-10<sup>12</sup> copies/mL | 5 (0.4%) | 8 (0.7%) |
| **Coexisting disease** |  |  |
| None | 547 (45.7%) | 480 (43.0%) |
| Cardiovascular disease | 178 (14.9%) | 130 (11.6%) |
| Respiratory disease | 47 (3.9%) | 64 (5.7%) |
| Metabolic disease | 94 (7.8%) | 99 (8.9%) |
| Nervous system disease | 170 (14.2%) | 170 (15.2%) |
| **Characteristics of clusters** |  |  |
| Number of days of exposure before enrollment, *median (IQR)* | 4.0 (3.0, 6.0) | 4.0 (3.0, 6.0) |
| Number of days of exposure before the intervention, *N (%)* |  |  |
| ≤3 days | 411 (34.3%) | 440 (39.4%) |
| 4-6 days | 668 (55.8%) | 551 (49.3%) |
| ≥7 days | 119 (9.9%) | 125 (11.2%) |
| Size of clusters, *median (IQR)* | 2.0 (1.0, 3.0) | 2.0 (1.0, 3.0) |
| Viral load of the index case, *n (%)* (N=549) |  |  |
| Undetectable (< 10<sup>4</sup> copies/mL) † | 47 (16.2%) | 42 (16.2%) |
| 10<sup>4</sup>-10<sup>6</sup> copies/mL | 85 (29.3%) | 68 (26.3%) |
| 10<sup>7</sup>-10<sup>9</sup> copies/mL | 125 (43.1%) | 129 (49.8%) |
| 10<sup>10</sup>-10<sup>12</sup> copies/mL | 33 (11.4%) | 20 (7.7%) |
| Type of contact with index case, *n (%)* |  |  |
| Household contact | 338 (28.2%) | 302 (27.1%) |
| Healthcare worker | 130 (10.9%) | 131 (11.7%) |
| Nursing home worker | 584 (48.7%) | 550 (49.3%) |
| Nursing home resident | 160 (13.4%) | 133 (11.9%) |
| Routine use of mask, *n (%)*‡ |  |  |
| Yes | 825 (68.9%) | 730 (65.4%) |
| No | 256 (21.4%) | 251 (22.5%) |
| NA | 117 (9.7%) | 135 (12.1%) |
| Sleeping in the same room as the index case, *n (%)* |  |  |
| Yes | 66 (5.51%) | 78 (6.99%) |
| No | 951 (79.4%) | 834 (74.7%) |
| NA | 181 (15.1%) | 204 (18.3%) |

**IQR:** interquartile range (i.e., 25<sup>th</sup> and 75<sup>th</sup> percentiles). **NA:** not available. **SD:** standard deviation.
\* Baseline PCR result was not available for 21 participants in the control arm and 14 participants in the intervention arm. † Pre-screening PCR was positive at the designated hospital lab prior to enrollment, but the result was negative (undetectable $< 10^4$ copies/mL) at the research lab from the swab collected on day 1. ‡ Routine use of mask refers to use at the time of exposure.

### PRIMARY OUTCOME

During the 14-day follow-up, 138 (6.0%) of 2,314 participants experienced a PCR-confirmed, symptomatic Covid-19 episode. The primary outcome was similar in the control arm (6.2%; 74/1,198) and the intervention arm (5.7%; 64/1,116; RR 0.89 [95% CI 0.54–1.46]) (Table 2). The incidence of each of the components of the primary outcome did not differ significantly between groups.

**Table 2.** Outcomes of hydroxychloroquine prophylaxis against Covid-19 (intention-to-treat population).

|  | **Control**<br>**arm**<br>**Events (%)** | **Intervention**<br>**arm**<br>**Events (%)** | **RR\* (95% CI)** |
| --- | --- | --- | --- |
| **Primary outcome** | N=1198 | N=1116 |  |
| **Overall (N = 2,314)** |  |  |  |
| PCR confirmed symptomatic Covid19 | 74 (6.2%) | 64 (5.7%) | 0.89 (0.54, 1.46) |
| Clinical and laboratory criteria | 60 (5.0%) | 49 (4.4%) |  |
| Hospital or vital records criteria | 14 (1.2%) | 15 (1.3%) |  |
| **PCR (-) at baseline (N =2000)** | N=1042 | N=958 |  |
| PCR-confirmed symptomatic Covid19 | 45 (4.3%) | 29 (3.0%) | 1.45 (0.73, 2.88) |
| Clinical and laboratory criteria | 37 (3.6%) | 24 (2.5%) |  |
| Hospital or vital records criteria | 8 (0.8%) | 5 (0.5%) |  |
| **PCR (+) at baseline (N=314)** | N=156 | N=158 |  |
| PCR-confirmed symptomatic Covid19 | 29 (18.6%) | 35 (22.2%) | 0.96 (0.58, 1.58) |
| Clinical and laboratory criteria | 23 (14.7%) | 25 (15.8%) |  |
| Hospital or vital records criteria | 6 (3.9%) | 10 (6.3%) |  |
| **Secondary outcomes (N= 2,000) †** | N=1042 | N=958 |  |
| Covid19 either symptomatically<br>compatible or PCR positivity<br>regardless of symptoms | 185 (17.8%) | 179 (18.7%) | 1.04 (0.77, 1.41) |
| Laboratory criteria ‡ | 67 (6.4%) | 58 (6.1%) |  |
| Clinical criteria □ | 150 (14.4%) | 144 (15.0%) |  |
| Hospital or vital records criteria | 8 (9.7%) | 5 (0.5%) |  |
| Serology positivity on day 14 | 91 (8.7%) | 137 (14.3%) | 1.6 (0.96, 2.69) |
| IgM positivity | 70 (6.7%) | 100 (10.4%) |  |
| IgG positivity | 82 (7.9%) | 118 (12.3%) |  |

**RR:** Risk ratio. **CI:** confidence interval.
\* Risk ratios are adjusted for contact-level variables (age, gender, region, and time of exposure).
† Excluding PCR positive at baseline.
‡ PCR confirmed either symptomatic or asymptomatic.
□ Symptoms compatible with Covid-19 regardless of PCR result
The components of the primary and secondary outcomes are not mutually exclusive.

Overall, the incidence of confirmed Covid-19 was higher in participants who tested positive in the baseline PCR (Table 2); 3.4% (74 of 2,000) participants with a negative PCR at baseline and 21.9% (61 of 279) participants with a positive PCR at baseline met the primary outcome criteria. The intervention was ineffective, regardless of the PCR result at baseline.

We observed an overall increased risk of Covid-19 with increasing viral load of the participant at baseline (Fig. 2A) and increasing viral load of the index case (Fig. 2B). The viral load of contacts who developed confirmed Covid-19 increased 4 log_10_ copies/mL throughout the follow-up, whereas that of contacts without Covid-19 remained unchanged (Fig. 2C). Pre-specified subgroup analysis of the primary outcome did not reveal between-group differences in the risk of Covid-19 according to the viral load of the participant at baseline, the viral load of the index case, the length of exposure, or the place of contact (Fig. 3).

**Figure 2.**
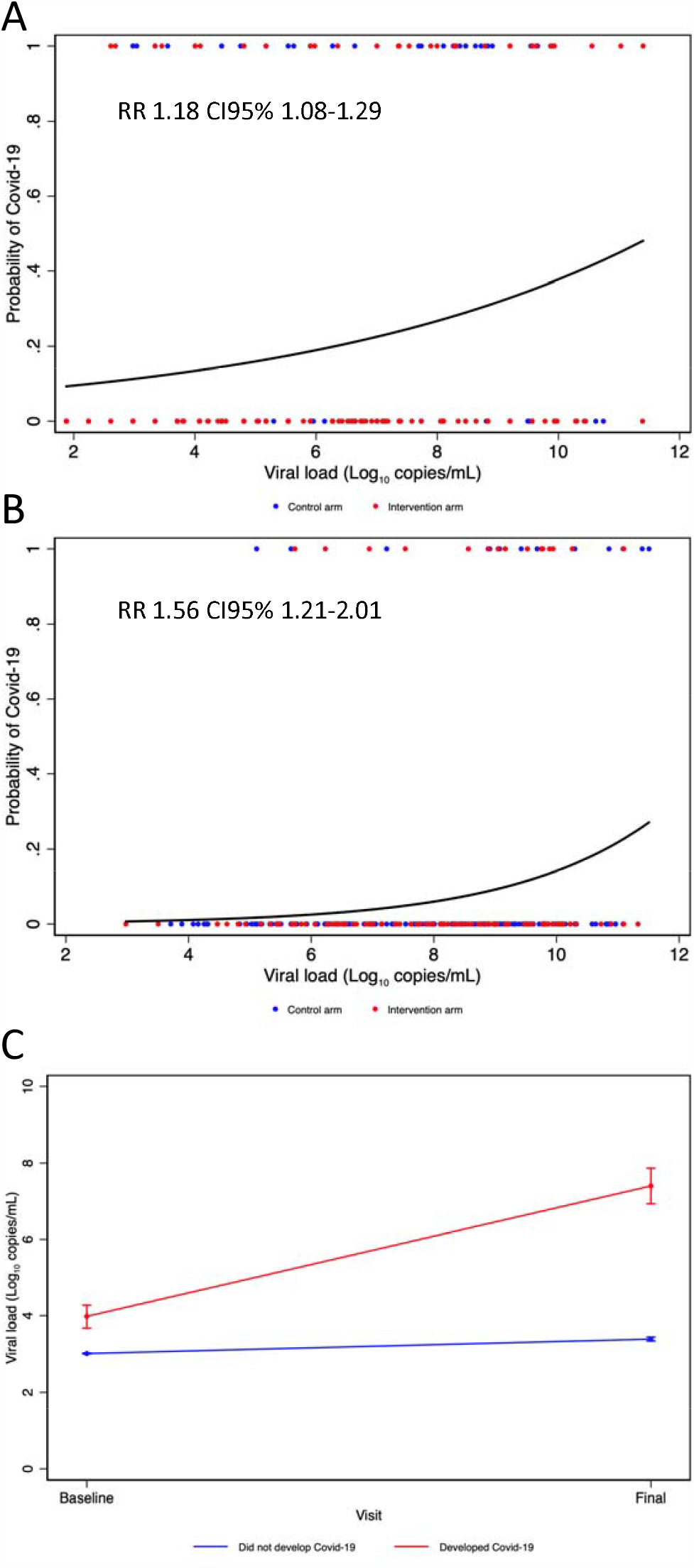
Association of baseline viral load of participants and viral load of their index case with breakthrough Covid-19 (ITT population) Panels A and B show the association of the participant’s viral load at baseline (A) and viral load of the index case (B) with the likelihood of developing PCR-confirmed symptomatic Covid-19 in the overall intention-to-treat population (aggregated data for the control and intervention arms). The dots are participants with (=1) or without (=0) the primary outcome of PCR-confirmed Covid-19. Panel C shows the viral load increase from baseline in participants who developed or did not develop Covid-19 (details are provided in Table S2, Supplementary Appendix).

**Figure 3.**
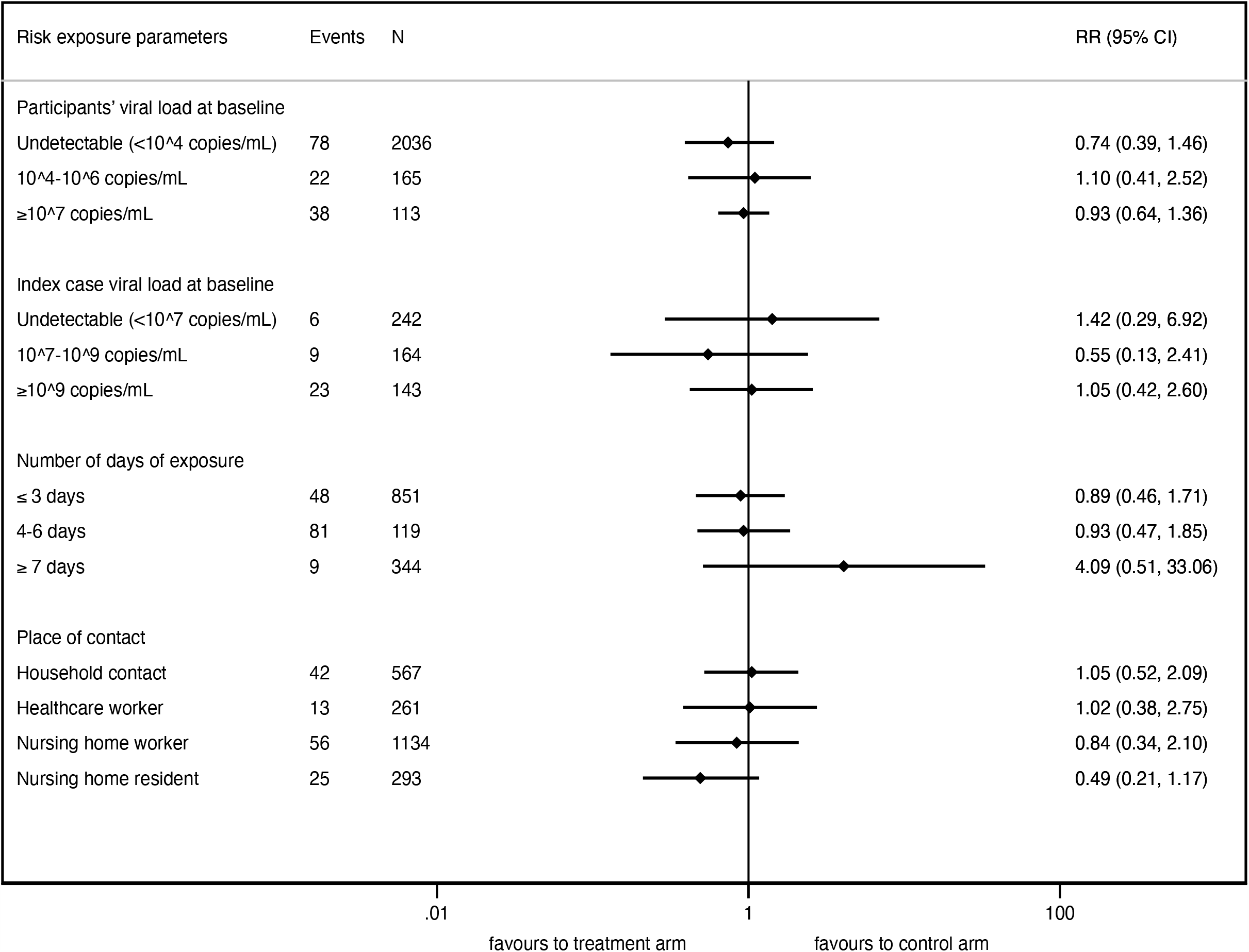
Subgroup analyses for the primary outcome according to risk of exposure factors (ITT population)

The survival analysis of the time to the primary outcome showed similar patterns in the two arms regarding confirmed Covid-19 onset from enrolment (median 14.0 vs. 14,0 days in the control and intervention arms, respectively; HR 0.9 [95%CI 0.6–1.5]) and from exposure (median 18.0 vs. 18.0 days; HR 1.0 [0.6–1.6]) (Fig. S3).

### SECONDARY OUTCOMES

Of the 2,000 participants who tested negative for SARS-CoV-2 in the baseline PCR, 364 (18.2%) either became PCR positive or developed symptoms compatible with Covid-19 throughout the follow-up period (secondary outcome, Table 2), without differences between study arms (17.8%, 185/1,042 control vs. 18.7%, 179/958 intervention; RR 1.04 [95%CI 0.77 1.41]). The virus-specific IgG/IgM positivity was higher in the intervention arm than in the control arm (6.7%, 70/1,042 control vs. 10.4% 100/958). Of 125 participants who became PCR-positive during follow-up, 30 (24.0%) were seropositive on day 14 (Fig S4).

### ADHERENCE AND SAFETY

Full adherence for the trial intervention was 97.5% (1,268 of 1,300) in the control arm and 95.1% (1,138 of 1,1197) in the intervention arm. In the safety population, 77/1,300 (5.9%) participants in the control arm and 671/1,197 (51.6%) in the intervention arm experienced at least one AE during 14 days of follow-up (Table 3). The most frequent treatment-related AEs among participants given HCQ were gastrointestinal (diarrhea, nausea, and abdominal pain) and nervous system disorders (drowsiness, headache, and metallic taste) (Tables S4). Thirty-one SAE were reported, 17 in the control arm and 14 in the intervention arm, none of them related to HCQ (Table S5). Six AEs of special interest were observed, including five episodes of self-limited palpitations potentially related to treatment (Table S6). Relevant safety data listings are provided in the Supplementary Appendix.

**Table 3.** No. of subjects experiencing at least one AE (Safety population).

|  | Control arm<br>N=1,300 | Intervention arm<br>N=1,197 | P-value |
| --- | --- | --- | --- |
| Reported full adherence to trial intervention | 1,268 (97.5%) | 1,138 (95.1%) |  |
| Adverse events |  |  |  |
| Any AE | 77 (5.9%) | 671 (51.6%) | <0.001 |
| Cardiac disorder (palpitations) | 1 (0.1%) | 5 (0.4%) |  |
| Gastrointestinal disorder (diarrhea, abdominal pain, and vomiting) | 33 (2.5%) | 510 (42.6%) |  |
| Nervous system disorder (headache, taste change, dizziness) | 32 (2.5%) | 260 (21.7%) |  |
| General disorder (myalgia, fatigue, malaise) | 10 (0.8%) | 103 (8.6%) |  |
| Intensity |  |  | <0.001* |
| Grade 1 | 44 (3.4%) | 573 (44.1%) |  |
| Grade 2 | 14 (1.1%) | 68 (5.2%) |  |
| Grade 3 | 2 (0.2%) | 13 (1.0%) |  |
| Grade 4 | 10 (0.8%) | 11 (0.8%) |  |
| Grade 5 | 7 (0.5%) | 6 (0.5%) |  |
| Serious AE † | 17 | 14 |  |
| Hospitalization | 12 | 11 |  |
| Deaths | 8 | 5 |  |
| Treatment-related Serious AE | 0 | 0 |  |
| AE of special interest (cardiac) ‡ | 1 | 5 |  |

\* overall p-value for grading
† None of the serious adverse events (SAE) were adjudicated as related to HCQ by the pharmacovigilance consultants.
Death and hospitalization were not mutually exclusive; five deaths occurred at the hospital while other participants died at a nursing home.
‡ Cardiac disorders were all palpitations episodes; 3 of 5 events in the intervention arm were adjudicated as possibly related to the study drug by the independent pharmacovigilance consultants. Details are provided in Table S6 (Supplementary material).

## DISCUSSION

Postexposure prophylaxis with HCQ did not prevent Covid-19 disease or SARS-CoV-2 infection in asymptomatic contacts exposed to a PCR-positive index case. In our cohort, the overall attack rate for the PCR-confirmed symptomatic Covid-19 was 6.0%, excluding subjects that were not enrolled because had symptoms before the baseline assessment. HCQ did not decrease the incidence of confirmed Covid-19 disease among contacts (6.2 vs. 5.7%). Our trial tested two possible effects of postexposure therapy: prophylaxis in contacts with negative PCR at baseline, and preemptive therapy in contacts with positive PCR at baseline (i.e., prevent progression of asymptomatic infection to disease). This dual scenario mirrors a real-life setting, where the PCR result of people exposed to a known Covid-19 case is usually not available immediately. Among PCR positive contacts at baseline (12% of subjects), the intervention had no apparent efficacy as early preemptive therapy. Of note a baseline positive PCR result significantly increased the risk of developing Covid-19 in our cohort, but a high proportion of participants with this lab result (79%) did not go on to develop symptomatic disease, thus reinforcing the need to quarantine or to increase testing of contacts even if asymptomatic. Also, of importance to the public health decision-making is that high Covid-19 viral load (>10^8^ log_10_ copies/mL (SD) results in more risk of transmission to contacts.

The intervention also did not reduce the transmission of SARS-CoV-2 (17.8% vs. 18.7%) or incidence of seropositivity. Notably, the overlap of positive PCR and positive serology was low, which could be related to both, the reported low rate of seroconversion in asymptomatic contacts^21^ or the higher risk of false negative PCR result on the initial stage of infection.^15^ Regarding safety, we observed a higher incidence of AE in the treatment group, albeit with low severity. This is an open-label study where the psychological components in the treated group cannot be excluded. Furthermore, the side effects reported were mainly at gastrointestinal level, while only five (0.3%) out of 1,479 events could be considered cardiac, thus not confirming previously published data that raised safety concerns.^22^ The safety results need to be interpreted considering the dose used, length of treatment, and the lack of ECG monitoring in the study.

The strengths of this study are the use of PCR and viral load titration at baseline, at day 14, and potentially when ill, and the measurement of viral load of the source index case to estimate risk of transmission. In addition, we included elderly persons (e.g. ages >90 years of age) from nursing homes. The study has some limitations. Unlike the common procedure in clinical trials, the informed consent signature took place after cluster-randomization. Nevertheless, allocation was revealed to participants after consent signature, therefore we believe the allocation concealment strategy was appropriate to prevent study participants from choosing to participate or not to participate. Owing to the urgency, the trial could not be masked with a placebo, which affected the rate of AE declared (AEs are not commonly reported in the control, non-placebo group), but it did not affect the attrition numbers in the control arm. However, it is worth mentioning that the laboratory staff who performed PCR tests remained unaware of the allocation of each sample.

Despite the promising in-vitro results that placed HCQ among the leading candidates for Covid-19 treatment and prophylaxis, ^23–25^ to date there is no strong argument to suggest that HCQ is effective. We provide high-quality evidence on the lack of efficacy of postexposure prophylaxis therapy with HCQ to prevent Covid-19 disease or SARS-CoV-2 infection. The data presented in this report is particularly valuable for the scientific community and policymakers involved in controlling the pandemic at the population level. Our findings encourage directing efforts to other antiviral candidates for postexposure prophylaxis.

## Data Availability

The study protocol and raw data will be available for a year upon request to the corresponding author

## CONTRIBUTORS

OM, LB, BC, CV, RMV, JC, CGB, MVM conceived, designed and wrote the manuscript,

MU, AA, CS, MC, PA, CA, AET, PL, SN, AN, JP, CQ, FMV, NRM, AS, CS, GFM, AF, GC, NP, NN contributed to the recruitment, clinical care, and follow-up of patients,

CT, AT, CL, EM, JP JR, AS, JZ, EM, JRU, SS analyzed and managed data

JA, JMA, JC, RF, MF analyzed data and reviewed the manuscript

EB, PC, ERM, LR Did all laboratory tests

JM, MC, MS, SG directed and managed the planning and execution of the project All authors reviewed and approved the final version of the manuscript

## CONFLICTS OF INTEREST

We declare no conflicts of interest

ACKNOWLEDGMENTS

This study was mainly supported by the crowdfunding campaign JoEmCorono (https://www.yomecorono.com/) with the contribution of over 72,000 citizens and corporations. The study also received financial support from Laboratorios Rubió, Gebro Pharma, Zurich Seguros, SYNLAB Barcelona, and Generalitat de Catalunya. Laboratorios Rubió also contributed to the study with the required doses of hydroxychloroquine (Dolquine®).

We thank Gerard Carot-Sans, PhD, for providing medical writing support during the revisions of the subsequent manuscript drafts, Eric Ubals (Pierce AB) and Òscar Palao (Opentic) for website and database management, Óscar Camps and OpenArms Non-governtmental organization for nursing home operations, Anna Valentí and the human resources department of the Hospital Germans Trias i Pujol for telephone monitoring. We are very grateful to Marc Clotet and Natalia Sánchez who coordinated the JoEmCorono crowdfunding campaign. We thank the institutional review board of Hospital Germans Trias Pujol and the Spanish Agency of Medicines and Medical Devices (AEMPS) for their prompt action for consideration and approvals to the protocol.

